# Causal Machine Learning Analysis of All-Cause Mortality in Japanese Atomic-Bomb Survivors

**DOI:** 10.1101/2025.09.03.25335009

**Authors:** Igor Shuryak, Zhenqiu Liu, Eric Wang, Xiao Wu, Robert L. Ullrich, Alina V. Brenner, Munechika Misumi, David J. Brenner

**Author notes:** Correspondence to: Igor Shuryak, MD, PhD Center for Radiological Research, Columbia University Irving Medical Center, 630 West 168^th^ Street, New York, NY 10032.

## Abstract

The health consequences of ionizing radiation have long been studied, yet significant uncertainties remain, particularly at low doses. In particular, traditional dose-response models such as linear, linear-quadratic, threshold, or hormesis models, all impose specific assumptions about low-dose effects. In addition, while the goal of radiation epidemiological studies is ideally to uncover causal relationships between dose and health effects, most conventional data analysis techniques can only establish associations rather than causation. These limitations highlight the need for new analysis methodologies that can eliminate the need for *a priori* dose-response assumptions and can provide causal inferences more directly based on observational data. Causal Machine Learning (CML) is a new approach designed to uncover how changes in one variable directly influence another, and with these motivations, a CML approach was, for the first time, implemented here to analyze radiation epidemiological data – in this case all-cause mortality data from Japanese A-bomb survivors. Compared to more traditional parametric approaches for analyzing radiation epidemiological data such as Poisson regression, CML makes no *a priori* assumptions about dose-effect response shapes (*e.g.,* linearity or thresholds). Extensive validation and refutation tests indicated that the proposed CML methodology is robust and is not overly sensitive to unmeasured confounding and noise. At moderate to high radiation doses, the CML analysis supports a causal increase in mortality with radiation exposure, with a statistically significant positive average treatment effect (p = 0.014). By contrast, no statistically significant causal increase in all-cause mortality was detected at doses below 0.05 Gy (50 mGy). These conclusions were drawn after adjusting for all available key covariates including attained age, age at exposure, and sex. We emphasize that this CML-based approach is not designed to validate or disprove any particular dose-response model. Rather this approach represents a new potentially complementary approach that does not rely on *a priori* functional form assumptions.

## INTRODUCTION

While high doses of ionizing radiation have well-established mutagenic, carcinogenic, and cytotoxic effects, considerable uncertainty remains regarding the health risks of low radiation doses (*1,2*). In particular, the complex biological pathways underlying diseases like cancer and cardiovascular disease remain incompletely understood, complicating efforts to definitively attribute disease occurrence to radiation exposure (*3–5*). As a result, radiation risk estimation relies on probabilistic models, with population-level probabilities expressed as functions of radiation dose. Detecting increased disease risks from low-dose radiation exposures is particularly challenging due to the small perturbations these exposures potentially induce compared with background disease levels (*6–10*).

The use of traditional radiation dose-response models, such as Linear No-Threshold (LNT) or threshold or hormesis models introduce further uncertainties into low dose risk estimation by imposing rigid assumptions about dose-response relationships (*7,10–16*). At low doses, it remains challenging to definitively validate or refute any of these models, and these differing frameworks introduce model selection uncertainty, requiring researchers to choose among different dose-response assumptions or combine models in an ensemble approach (*17,18*).

As a consequence of these limitations, and despite the availability of a number of high-quality radiation epidemiological studies, risk estimates at low radiation doses remain very uncertain. Thus, for example, while the large cohorts of Japanese atomic bomb survivors - who were exposed to a wide range of radiation doses at high dose rates - have been extensively studied (*16,19,20*), the effects of low radiation doses in these cohorts still remain incompletely characterized (*21–23*). Similar uncertainties arise in populations exposed to more prolonged radiation exposure, such as nuclear industry workers (*3,24,25*).

In addition, while the goal of radiation epidemiological studies is ideally to uncover *causal* relationships between dose and health effects, most conventional data analysis techniques can only establish *associations* between dose and health effects, rather than assessing a causal link.

These limitations highlight the need for new analysis methodologies that can: a) eliminate the need for *a priori* dose-response assumptions, and b) provide causal inferences between radiation dose and risk, more directly based on observational data.

While establishing and quantifying causal effects is a difficult challenge, particularly based on observational data, its fundamental importance is fueling an increasing body of research and a growing toolkit of methodologies (*26–35*). Following from the pioneering work of Rubin (*36,37*) and Pearl (*26,27*) on causal inference, Causal Machine Learning (CML), for example with its implementation using double/debiased machine learning (DML) (*28,29,35*) and causal forests (CF) (*31,32*), has emerged as a particularly promising approach to address these challenges. Traditional predictive modeling techniques that are based on correlations are designed to predict what the outcome value (*e.g.,* the mortality rate) will be as function of all the predictor variables (*e.g.,* radiation dose and other covariates such as age and sex), but they are not designed to estimate how the outcome will change if a specific variable is modified. By contrast, causal inference is designed specifically to estimate what causal effect of intervening on a treatment or exposure variable will have on the outcome (*e.g.,* how will giving a larger dose of a medical treatment or being exposed to a higher radiation dose affect the mortality rate). CML extends causal inference to situations where the number of covariates can be large, interactions between them and the treatment and outcome can be complex, and the functional forms for these interactions can be nonlinear and *a priori* unknown. In other words, in contrast to traditional statistical and predictive ML methods, which focus on minimizing overall prediction errors rather than on the potential causal effects of a specific treatment or exposure variable like radiation, CML treats the causal variable as conceptually distinct from other features which may affect the outcome. This distinction is important because traditional predictive models may prioritize strong predictors like age and sex over the treatment effect, potentially distorting or ignoring it during regularization. So, unlike conventional ML, which excels in prediction but fails to distinguish correlation from causation (*27,38*), CML is designed to explicitly estimate causal relationships by modeling treatment effects (in this case radiation dose) while accounting for confounding variables. This capability has the potential to be highly advantageous in radiation epidemiology, where understanding how radiation exposure causally influences health outcomes across different populations is the desired goal.

With these motivations, a modern CML approach has been implemented here to analyze radiation epidemiological data – in this first example all-cause mortality data from Japanese atomic bomb survivors. To our knowledge, this is the first use of such a methodology in radiation epidemiology. Our goal is to outline a flexible, data-driven perspective on radiation risk estimation, which can be used for causal inference without imposing any rigid parametric assumptions on the dose-response shape, or on interactions of radiation with other factors. Causal machine learning is a new approach for analyzing radiation epidemiological data, and this study demonstrates its potential to complement traditional approaches and particularly to advance our understanding of low-dose radiation effects.

## METHODS

### Causal Machine Learning (CML)

One of the main strengths of CML is its ability to perform causal inference on large data sets with multiple variables and complex interactions between them. Strict functional form assumptions about the causal effect are not required which, as discussed above, is an important issue for analyzing the effects of low radiation doses.

Double debiased machine learning (DML) – a state-of-the art implementation of causal machine learning – consists of two main stages: First estimation of nuisance functions: this includes a treatment model predicting radiation dose from covariates (deconfounding) and an outcome model predicting health outcomes from covariates (denoising). Second, ultimate causal effect estimation (*28,29*): here DML uses a doubly robust estimator to combine these components, allowing for the estimation of both average treatment effects (ATE) and conditional average treatment effects (CATE) at the individual or subgroup level. Importantly, this doubly robust property means that unbiased estimates can be obtained even if only one (but not necessarily both) of the nuisance function models is correctly specified. DML employs cross-fitting or sample splitting, which involves dividing datasets into multiple partitions, using some subsets to fit the nuisance functions and others to estimate treatment effects, thereby minimizing overfitting and enhancing generalizability.

The DML framework conveniently allows the researcher to use different model types, as appropriate for the data type and research question, for the two stages of the process. For example, various ML algorithms, such as random forests (*39*), XGBoost (*40*), and elastic net regression (*41*), can be applied within the DML framework at the nuisance function estimation stage. Importantly, flexibility in model type choice is also possible at the causal effect estimation stage and rigid response shape assumptions are not required. Here we used causal forests (CF), a tree-based ensemble method derived from random forests, for this purpose (*32*). CF are extremely flexible and stand out for their ability to capture heterogeneous treatment effects across subgroups (*31,32*).

### A-bomb Data and Data Pre-processing

We analyzed the publicly available Radiation Effects Research Foundation (RERF) mortality data set from “Studies of the Mortality of Atomic Bomb Survivors, Report 14”, which covers the period from 1950 to 2003 (*42*). RERF, along with its predecessor, the Atomic Bomb Casualty Commission (ABCC), has conducted the Life Span Study (LSS), a long-term mortality study initiated in 1950. The primary aim of the LSS is to assess the late health effects of ionizing radiation exposure resulting from the atomic bombings. The LSS cohort includes over 120,000 individuals comprising atomic bomb survivors from Hiroshima and Nagasaki (86,611 with radiation dose estimates), as well as 26,529 unexposed residents who were not in either city at the time of the bombings (*42*). The LSS Report 14 data set, which was the basis of our analysis, includes data for 86,611 in-city survivors grouped into a person-year and event table stratified by city (Hiroshima / Nagasaki), sex (male / female), age at exposure (15 categories), attained age (21 categories), follow-up period (11 categories), radiation dose (DS02 weighted colon dose, 22 categories), distance from the hypocenter (<3 km / 3-10 km) and Adult Health Study (AHS) participation (not participant / original AHS / AHS extension). The person-year table has 53,782 rows with total of 3,294,210 person-years and 50,620 deaths. The mean and median colon doses, weighted by person-years, were are 0.116 and 0.010 Gy, respectively. The majority of individuals received very low doses, with 38,509 exposed to < 0.005 Gy (5 mGy), 68,470 (or 79% among in-city survivors) exposed to <0.1 Gy, and only 2,387 and 624 individuals estimated to have received >1 Gy or >2 Gy, respectively (*42*). The distribution of doses across person-years is illustrated in **Fig. 1**. The cohort includes both males (35,687) and females (50,924), and individuals exposed at Hiroshima (58,494) and Nagasaki (28,117).

**FIG. 1:**
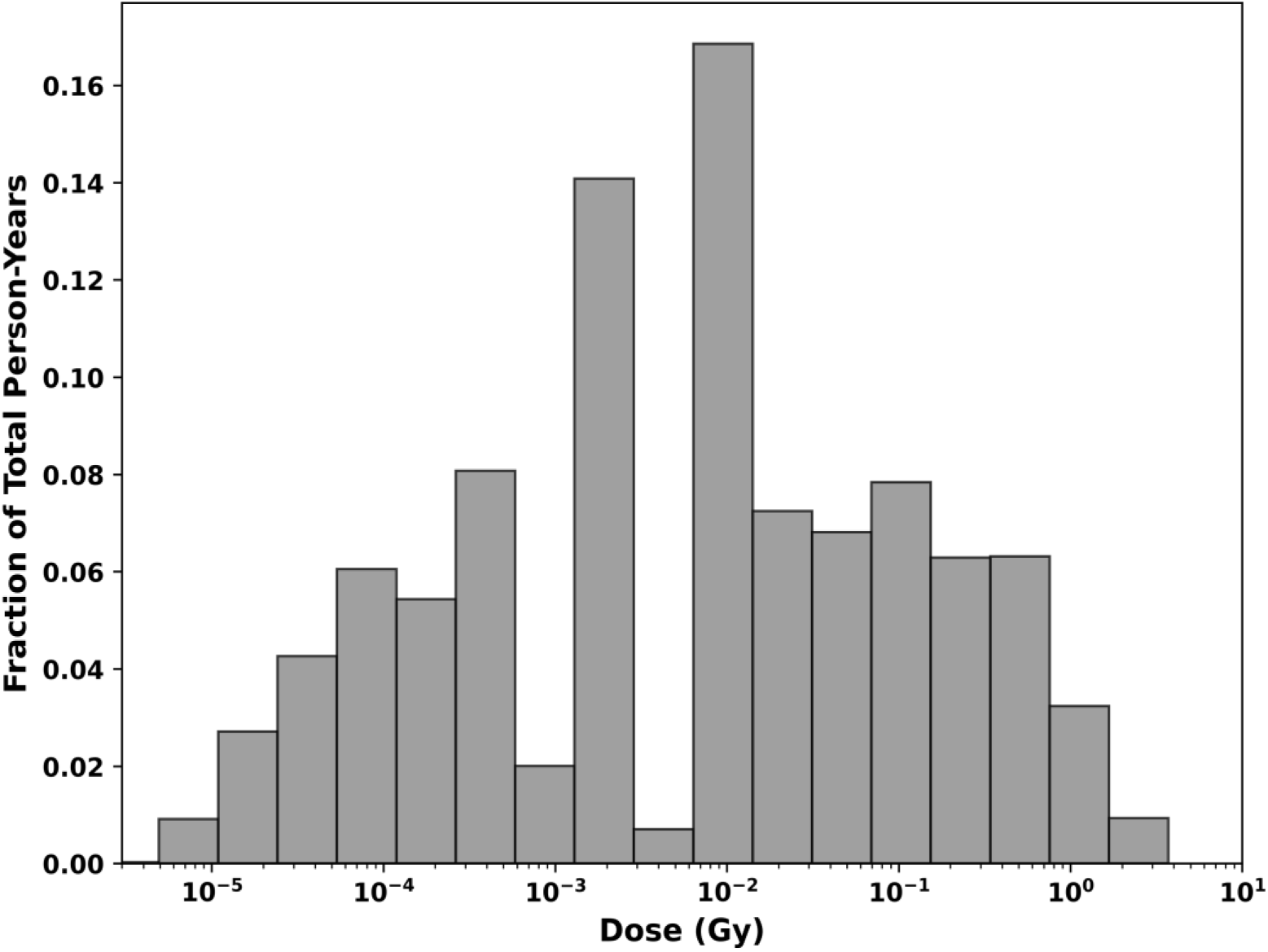
Illustrating the proportional distribution of person-years exposed to different doses in the data set analyzed. Individuals exposed to zero dose are not shown on this log dose scale.

Since the Report 14 data set provides only aggregated person-year tables (due to privacy constraints on individual data), all modeling, inference, and resampling procedures were conducted at the subgroup level.

The data set was downloaded on September 20, 2024 from the RERF website (*43*) and analyzed in the Python programming language. Categorical variables were recoded as binary indicators: of city, sex and Adult Health Study cohort membership. Continuous features like attained age and age at exposure were included in continuous form (not as categories), and the outcome variable was all-cause mortality rate.

The death event rate per person-year was transformed using the natural logarithm, expressed as *ln*(death/pyr). Because this transformation is undefined for zero-event samples, a standard K-means clustering algorithm (*44*) was used to merge these samples with similar samples that had at least one event. The clustering process considered radiation dose, city, sex, age at exposure, attained age, and AHS status as features. Within each cluster, weighted averages were calculated for each feature, using person-years as weights. The K-means clustering was implemented in the scikit-learn Python module (*45*), with a custom function for weighted averaging. These pre-processed data were then analyzed by CML, as described below.

### Causal Machine Learning (CML) Analysis

To estimate the causal effects of radiation exposure on, in this case, all-cause mortality, we applied Causal Forest DML algorithm from the EconML Python package (*46*). This doubly robust causal forest implementation employed specialized configurations for both outcome (log event rate) and treatment (dose) submodels to mitigate overfitting risks. Both outcome and dose used constrained random forest regressors with 50 trees, a maximum depth of 6 nodes, and regularization through minimum leaf/split thresholds (3 samples). Feature subsampling at 80% per split further reduced variance by decorrelating individual trees, while the modest tree count was chosen to balance computational efficiency with ensemble diversity. The double robustness property of this method reduces its sensitivity to model misspecification, which is particularly valuable in complex causal inference tasks with continuous treatment and outcome variables, as in this study. Sample weights proportional to the number of person-years of observation for each data sample were used to avoid biasing the analysis towards samples with small numbers of person years.

We used a polynomial featurizer in the CausalForestDML procedure to expand the treatment variable into second-order terms; this improved the capability of the DML to “learn” complex radiation response patterns. This approach does not imply any global assumptions about the dose-response shape of the analyzed causal effects - the causal forest retains full flexibility to estimate radiation effects of any shape.

The causal forest DML was fitted to a training dataset consisting of a randomly chosen 75% subset of the full clustered dataset, using 10-fold cross-validation and 10 Monte-Carlo iterations. The 10-fold cross-validation involves dividing the training data into 10 subsets. The model was trained on 9 of these subsets and validated on the remaining one, rotating through all 10 possibilities. This process helps to assess how well the model generalizes to unseen data and reduces overfitting. The 10 Monte Carlo iterations involve repeating the entire cross-validation process 10 times, each time with a different random split of the data into folds. This adds another layer of randomization and helps to account for the variability that can occur due to the specific way the data is split. The results from these repeated cross-validations are then averaged to provide a more stable and reliable estimate of the model’s performance and treatment effects. This approach helps to mitigate the impact of any particular random split of the data and provides a more robust assessment of the model’s capabilities. Following the training phase, causal effect estimates were calculated on a separate testing dataset, consisting of the remaining 25% of the data.

Uncertainties in the causal effects and corresponding confidence intervals were estimated based on the bag-of-little-bootstraps (BLB) approach (*47*). The BLB method creates multiple small subsets of the original dataset, performs bootstrap resampling within each subset, calculates desired statistics for each resample, and aggregates results across all subsets. This approach is computationally efficient while maintaining statistical robustness.

### Validation of the CML Methodology Using Simulated Data

The proposed analysis method of data clustering + causal modeling was tested using a variety of different simulated data sets, each derived from a different assumed underlying radiation dose-response shape. More details are provided in **Supplementary Information 1 (Validation Studies on Simulated Data)**. The purpose of these validation studies was to test the capability of the clustering + causal modeling procedure to recover the assumed dose response function. We tested two model configurations. The first configuration used raw dose values without transformation, relying solely on the causal forest to approximate non-linearities. The second incorporated polynomial features using the featurizer described above. The results of these analyses (see **Supplementary Information 1**) suggests that the polynomial featurizer accurately reconstructed the simulated dose-response, and this approach was thus used in the analysis of the A-bomb data.

### Refutation Tests and Sensitivity Analyses

For the actual A-bomb data, the true causal effect is, of course, unknown, so refutation tests and sensitivity analyses were used to assess the robustness of the causal effect estimates in response to data perturbations. These tests were designed to challenge the assumptions underlying the causal model and evaluate whether the estimated effects remain consistent under various perturbations or alternative scenarios. By attempting to falsify the causal conclusions, refutation tests help ensure that the results are not driven by unmeasured confounding, model misspecification, or data artifacts. In machine learning-based causal analyses, such as this one, refutation tests are particularly important because they provide a way to validate complex models that may rely on numerous assumptions. They also help identify potential vulnerabilities in the analysis, such as sensitivity to unobserved variables or spurious correlations. However, we acknowledge that the structure of our synthetic variables is relatively simple and differs somewhat from that of real-world confounding.

Details and results of these tests are described in **Supplementary Information 2 (Refutation Tests on RERF Data)**. In summary, the refutation analyses collectively indicated that the causal modeling strategy is quite robust to unmeasured confounding, noise, and spurious associations.

### Feature Importance Analyses using SHAP Values

SHAP (SHapley Additive exPlanations) values represent a powerful method for interpreting ML models by quantifying the contribution of each feature to individual predictions based on game theory principles (*48*). SHAP values are calculated by considering all possible feature combinations and determining each feature’s average marginal contribution to the model’s prediction. In the context of this causal model, SHAP values help explain how different features influence the heterogeneity of treatment effects estimated by the model. This analysis provides insights into which covariates are most important in determining how the treatment effect varies across the population.

We used the Kernel SHAP approach (*49*) to compute SHAP values. This approach is particularly useful when dealing - as here - with complex models that lack built-in exact SHAP implementations. The approach does, however, have some limitations such as assuming feature independence (*49*), which may not hold in reality. For this reason, we calculated the Pearson correlation matrix of all the features in the model and used these to guide our interpretation of the SHAP values.

## RESULTS

### Validation of the CML Methodology using Simulated Data

In order to validate our CML methodology before applying it to the A-bomb survivor data, the goal of these data simulations was to assess the capability of the CML approach to reconstruct different simulated dose-response data sets. Detailed results are described in **Supplementary Information 1**, using the proposed methodology (*ln-*transformation of the event rate and data clustering followed by CML analysis) on simulated data sets with different dose response shapes and magnitudes.

In summary, the CML methodology performed well on the simulated data sets. Performance was best when the event rate was relatively high, with a high baseline intercept and/or an increase with increasing dose. As expected, performance was somewhat worse in cases when the event rate was low due to a low baseline and/or a steep negative effect of radiation dose. Overall, for plausible radiation dose-response scenarios the proposed methodology performed well, and the modeling procedure was able to reconstruct the simulated radiation dose-response patterns.

### CML Analysis of RERF Mortality Data

After evaluating simulated data sets, the proposed analysis pipeline was applied to the RERF all-cause mortality data set consisting (see above) of 39,129 rows with zero deaths and 14,653 rows with non-zero deaths. The iterative K-means clustering algorithm described above condensed these data into 1,201 clusters, where each cluster is a subgroup of individuals with similar covariates (*e.g.,* age at exposure, sex, city, attained age). The distributions of the main features of the clustered data set was very similar to those in the original data set: For example, in the original data set mean and standard deviation for age at exposure were 27.8 ± 17.9 years and the corresponding values in the clustered data set were 30.0 ± 18.0; for attained age the numbers were 57.3 ± 20.1 and 60.5 ± 18.5, respectively. The Pearson correlation matrix of features in this clustered data set is shown in **Fig. 2**.

**FIG. 2.**
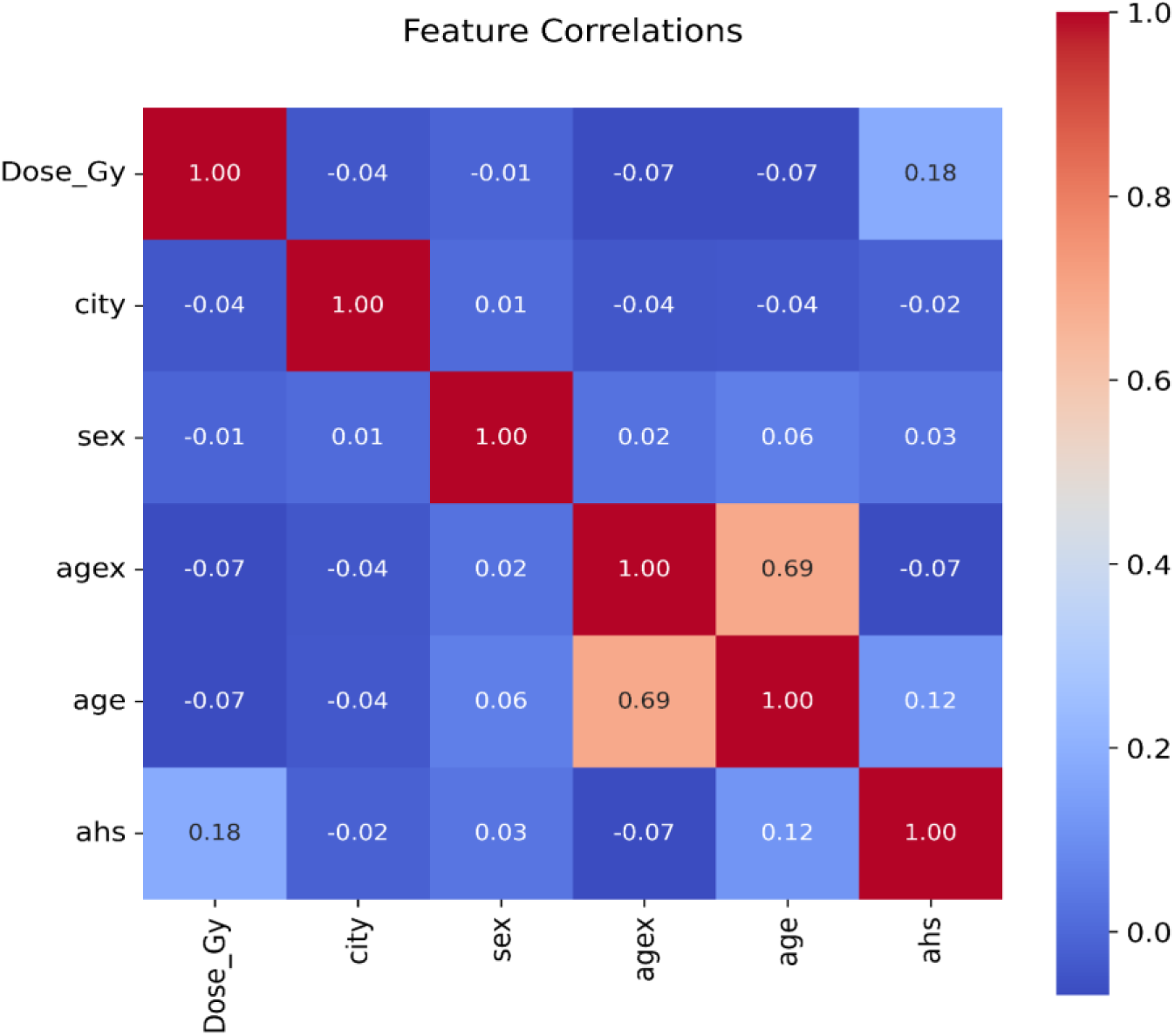
Pearson correlation matrix of features in the clustered all-cause mortality data set. Correlation coefficients are listed in each cell and color coded for easier visualization.

This CML approach was used to estimate the Relative Risk (RR) of mortality within the clustered data set. (**Fig. 3**). As expected, at medium and high radiation doses the derived RRs suggest a causal increase in radiation risk with increasing dose (**Fig. 3A**). Averaged over all doses, the Average Treatment Effect (ATE), on a scale of log-transformed mortality rate, was 0.30 Gy^-1^ (95% CI: 0.06 - 0.53, p-value 0.014). This value is consistent with the corresponding sex-averaged estimate from an earlier analysis of overall mortality by Ozasa *et al.* (*42*) (0.22 Gy^-1^, 95% CI: 0.18 – 0.26), though of course the methodology used in the current analysis is conceptually different from the earlier parametrically-based analysis by Ozasa *et al.* (*42*).

**FIG. 3.**
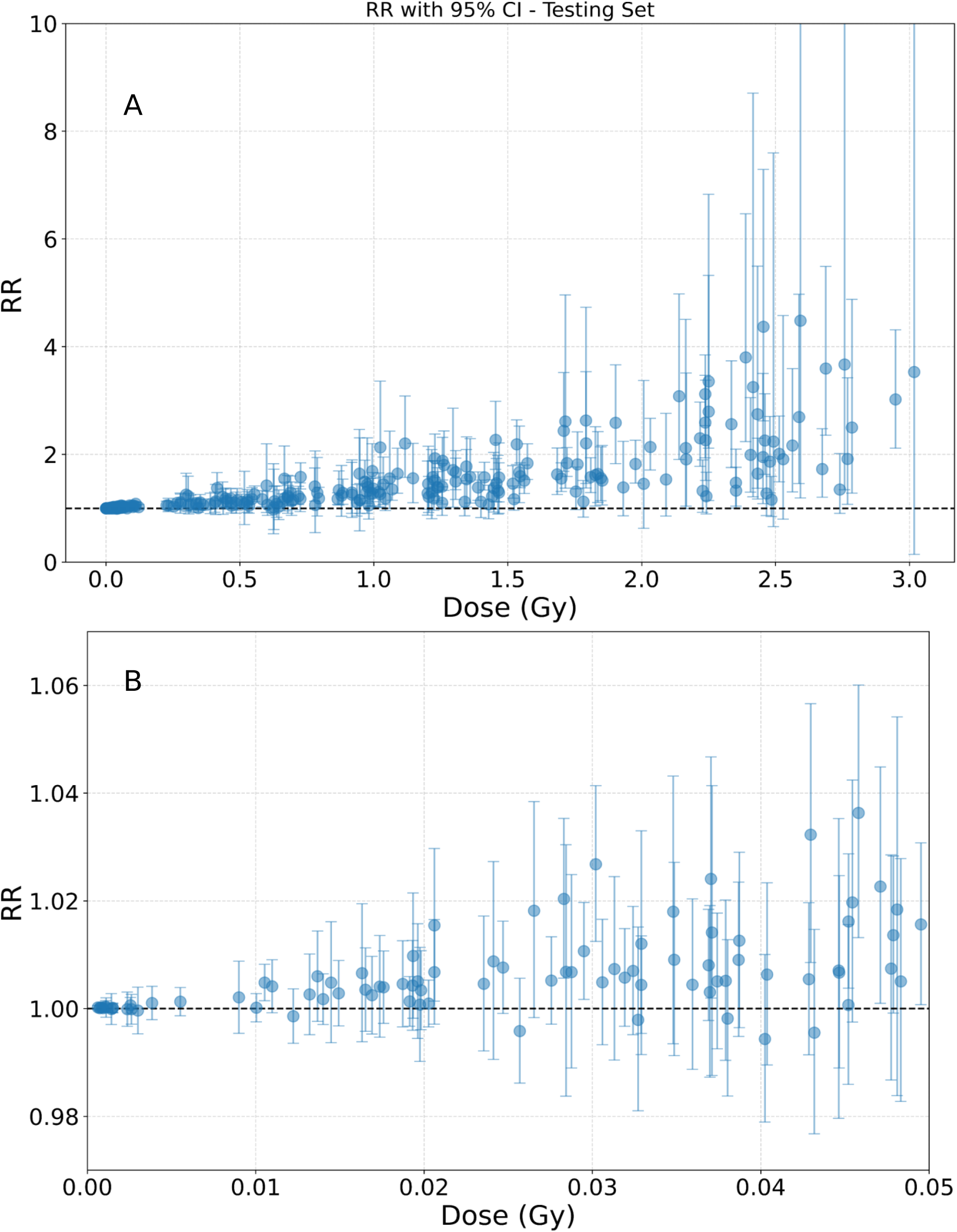
Estimated radiation-induced relative risks (RRs) for all-cause mortality for each of the samples (data clusters) in the testing data set. Error bars represent 95% CIs. Top Panel (A): Full dose range; Bottom Panel (B): Magnified view of low-dose (<0.05 Gy) region.

At low radiation doses, the derived Relative Risks (**Fig. 3B**) were generally close to 1 with confidence intervals including 1, suggesting no detectable causal radiation effects in this low dose range. For example, and motivated by the study by Grant *et al.* (*22*) where a significant dose response was observed for solid cancer incidence in the dose range from 0 to 0.1 Gy (100 mGy) but not in lower dose ranges, we repeated our CML-based analyses but with doses restricted to below 0.05 Gy (50 mGy); in this dose range the Average Treatment Effect (ATE) was not significantly different from zero: −0.23 Gy^−1^ (95% CI: −5.77 −5.32, p-value 0.936).

### Effect Modifiers: SHAP Value Analyses

SHAP value analyses, intended to identify key modifiers of the estimated radiation effect, are summarized in **Fig. 4**. Age at exposure and attained age contributed the most (**Fig. 4 A-B**), which is consistent with previous epidemiological work on RERF data sets (*19–22,42*). Their SHAP values tended to decrease with increasing values for both features, suggesting that the strongest radiation effect was estimated for those exposed in childhood and who died at a relatively young attained age, whereas older age at exposure and/or older attained age were associated with reduced radiation effects on mortality (**Fig. 4 C-D**). These patterns are in line with expectations.

**FIG. 4.**
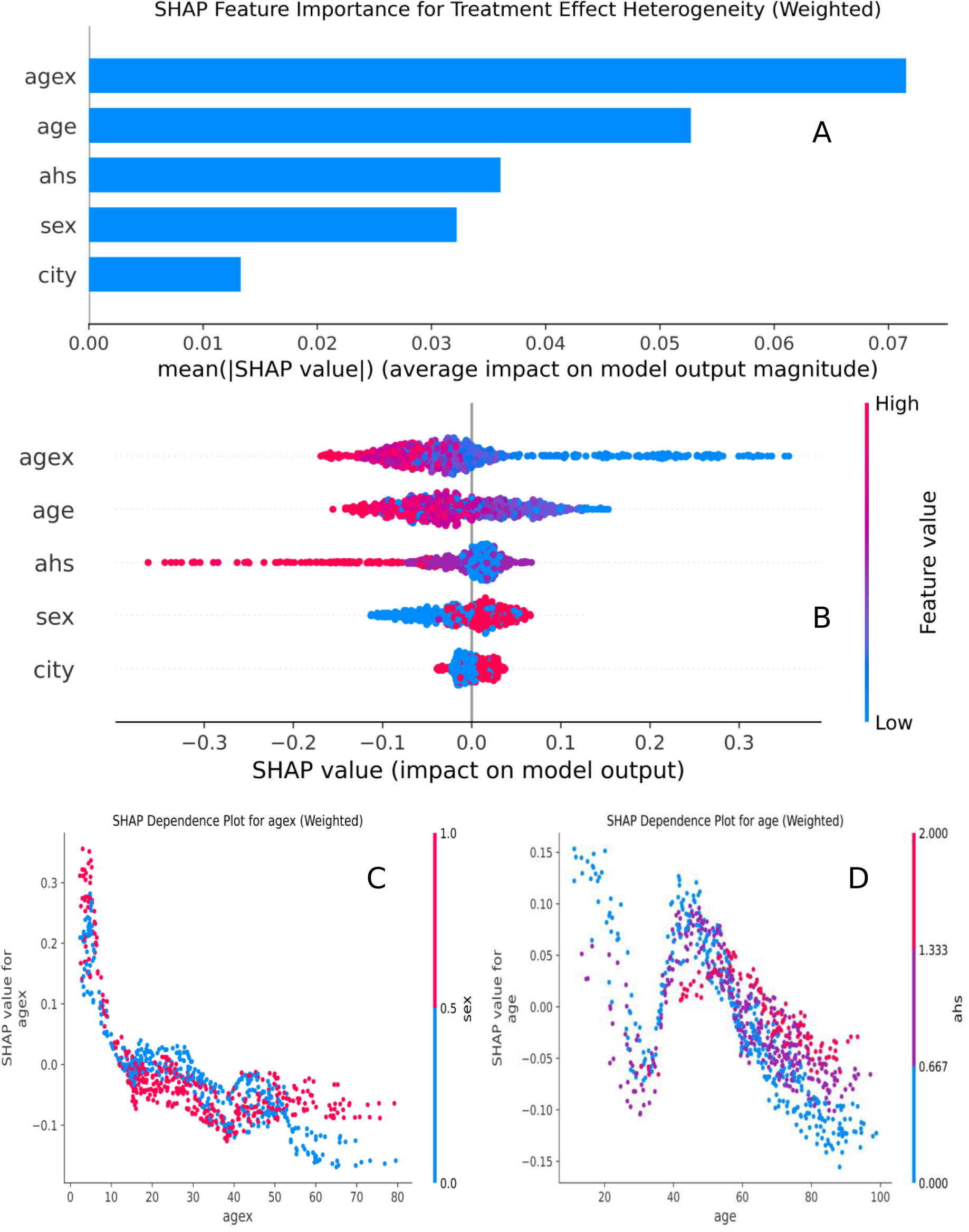
SHAP value analysis for all-cause mortality. Panel A = Bar graph showing the ranking of features by strength of contribution to the causal model, based on mean absolute SHAP values. Panel B = More detailed sample-level view of SHAP values for all features. Here each circle represents a sample from the training data set, and the blue to red color scale represents the magnitude of each feature. Negative SHAP values indicate that the particular feature reduces the radiation effect for the particular sample, positive values indicate that the feature increases the radiation effect, and zero indicates no influence on the radiation effect. Panels C and D = Sample-level SHAP values for continuous features age at exposure and attained age as function of feature values.

As discussed above, the kernel SHAP explainer methodology used in this causal model can be sensitive to strong correlations between features (*49*). In this analysis attained age and age at exposure were indeed strongly correlated, as shown by the Pearson correlation matrix (**Fig. 2**). For this reason, the contributions of attained age and age at exposure may potentially not be reliably disentangled from one another, whereas the other features were only weakly correlated and their approximate SHAP values should be more reliable.

As illustrated in **Fig. 4**, participation in the AHS study also made an appreciable contribution, with somewhat lower radiation risks for AHS participants *vs*. non-participants. The effect of sex suggested somewhat higher radiation risks for females than for males. City (Hiroshima vs Nagasaki) made the smallest contribution among all the available features.

### Sensitivity Analyses and Refutation Studies

Detailed descriptions and results of the sensitivity analyses and refutation studies are provided in **Supplementary Information 2**. Briefly, the results of the different refutation tests described there suggest that the causal modeling approach implemented here produces results that are robust to realistic magnitudes of unmeasured confounders and noise.

## DISCUSSION

The health effects of ionizing radiation have been studied for over a century, yet significant uncertainties and controversies persist in estimating radiation-induced health risks, particularly at low doses, where any radiation risks are much smaller than background risks.

Two of the main issues are, first, the use of traditional parametric radiation dose-response models, such as LNT, threshold or hormesis models; assuming such models complicate risk estimation by imposing *a priori* assumptions about the shape of the dose-response relationships. In addition, whilst the desired goal of epidemiological studies is to uncover causal relationships, most conventional analysis techniques are designed to detect correlations and associations between dose and risk, rather than assess causation.

Modern Causal Machine Learning (CML) approaches, based on the principles of causal inference theory combined with machine learning algorithms (*26,29,32,37*), offer promising solutions to these issues, and we describe here a first application of this approach to radiation risk estimation. Specifically, causal inference is designed to uncover how changes in one variable directly influence the outcome - the variables in our case being radiation dose and health consequences.

CML methods can implement causal inference principles in high-dimensional datasets and detect nonlinear interactions among variables such as demographics, genetics, and lifestyle factors (when such data are available). Importantly, they have the potential to flexibly adapt to data patterns without the need for strict parametric constraints. For example, the causal forest DML method used here does not require any *a priori* specification of how radiation dose can affect the mortality rate, or how covariates like age and sex can interact with this radiation effect – all these dependences are modeled by the flexible forest-based methods which consist of ensembles of decision trees.

Before using this CML approach to analyze “real-world” data, in this case from atomic bomb survivors, we performed an extensive series of validation studies to assess the utility of this CML approach for assessing the causal effects of radiation exposure. These validation studies used simulated data generated from differently-shaped assumed dose responses. The tests suggested that the approach was robust: it resisted synthetic confounders unless they very strongly affected both treatment and outcome, was insensitive to realistic amounts of noise, and correctly detected no effect when treatment was shuffled, all confirming that the model is capable of identifying true causal effects.

We then used this CML approach to analyze a large-scale data set of mortality among atomic bomb survivors, with estimates adjusted for key covariates including attained age, age at exposure, and sex. Our findings at moderate to high radiation doses were broadly as expected, indicating a causal increase in mortality with increasing radiation dose (**Fig. 3 A**). However, the CML analysis did not support a significant causal relationship between radiation dose and overall all-cause mortality at very low doses. For example, below 0.05 Gy (50 mGy), the estimated Relative Risks were close to 1 with 95% confidence intervals that largely encompassed the null (**Fig. 3B**), and if the CML analysis was restricted to only this low dose region the Average Treatment Effect (ATE) was not significantly different from zero (p-value 0.936).

Causal effect heterogeneity was further explored using SHAP values (**Fig. 4**). As expected, the analysis identified age at exposure and attained as the strongest modifiers of radiation risk, followed by AHS study participation, sex and city. While previous studies used parametric modeling to capture the relationships and interactions between these covariates and the radiation effect, the current CML study did not rely on such assumptions. Thus the shapes of the SHAP dependences (**Fig. 4 C-D**) were not constrained by any specific functional forms, potentially revealing subtleties that parametric modeling could overlook.

This first approach to a causal analysis of radiation epidemiological data has, of course, some limitations, particularly related to the structure of the available data. Specifically, the available A-bomb survivor dataset is structured in a person-year table format (rather than individual person data), which necessitated use of a clustering data preprocessing approach and analysis at the subgroup level rather than at the individual level. While the data set that was analyzed still contains a fair degree of data granularity (1,201 data cluster points analyzed, describing 86,611 individuals), this clustering procedure undoubtedly results in loss of information and some bias and noise - though simulation studies suggested that such effects are not overwhelming. In particular our bootstrap confidence intervals estimation may somewhat underestimate uncertainty by not capturing within-subgroup variation. It is emphasized that this issue is related to the structure of the RERF data set, and would not arise if/when individual person data becomes available – as is the case for other relevant radiation epidemiology data bases, such as the CEDR nuclear worker data sets (*50*).

A more general issue is that if important confounding variables are unobserved or omitted in the primary data, even advanced CML techniques cannot completely eliminate the potential for bias. As described above, to assess the potential impact of unmeasured confounding, we conducted extensive refutation testing which indicated relatively low sensitivity to such confounding in this analysis. That said, some potentially important variables — such as tobacco use, lifestyle factors, and genetic / molecular markers — were not captured in this analysis. As additional covariate data become available, the CML framework can readily incorporate them; causal forest models, in particular, are well-suited to handling large and complex feature sets.

In conclusion, this study illustrates how causal inference, combined with ML methods, can supplement traditional analysis approaches in analyzing radiation epidemiological data. By avoiding strict dose-response assumptions and leveraging flexible, data-driven causal-based techniques, CML provides a powerful framework for advancing radiation risk assessment, including at low doses where there is still much uncertainty. We emphasize that this CML-based approach is not designed to validate or disprove any particular dose-response model. Rather this approach represents a new complementary approach which does not assuming a specific dose-response function. Future work can extend this approach to other irradiated populations for which large-scale data sets exist, such as nuclear workers who received prolonged low-dose radiation exposures (*50*), as well as medically exposed populations.

## Supplementary Information

Supplementary Information 1: Validation Studies on Simulated Data.

Supplementary Information 2: Sensitivity Analyses and Refutation Tests using the RERF data set.

## SUPPLEMENTARY INFORMATION

### Supplementary Information 1: Validation Studies on Simulated Data

#### Data Simulation

Before applying the CML approach described in the main text to the real RERF mortality data, detailed simulation studies were conducted to test the ability of this method to perform when the causal effect’s magnitude and shape were known. Simulated mortality data was generated using a Poisson regression model using the following equation, where *D* is radiation dose, β values are model parameters, pyr denotes person-years, age, agex, sex and city are features from the real data set, and λ is the simulated mean death count for each data set sample (row):

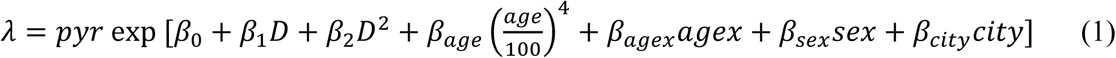

Based on this equation, the death counts column in the mortality data set was replaced with a random Poisson vector with mean λ. All other columns in the data remained unperturbed. Thus, the covariates data set structure was untouched, but the real outcome was replaced with a simulated one. Multiple simulations of this semi-synthetic approach with different β values were generated and analyzed.

#### Poisson Regression on Clustered Simulated Data

After clustering of the simulated data, we performed Poisson regression to recover the coefficients of the model used in data simulation. The same model structure was used as in the simulation model (Eq. 1), but coefficients were now unknown and adjustable, intended to be recovered by Poisson regression. The purpose of this step was to serve as a validation of the clustering process to estimate whether or not it preserves the main data set properties and that the coefficients of the simulation model can be reconstructed accurately.

#### Simulated Data: Analysis Results

We found that if the number of K-means clusters in the iterative procedure was >1000, Poisson regression performed well at recovering the parameters of the simulation model. The results of 500 simulations are summarized in **Fig. S1** to illustrate this. For example, when the parameter values during simulation were β_0_ = −2, β_1_ = 1, β_2_ = −0.25, β_age_ = 1, β_agex_ = −0.02, β_sex_ = 0.5, and β_city_ = 0, the corresponding fitted values on clustered data (1500 clusters) with their standard errors were very similar to the simulated values: β_0_ = −1.996 ± 0.003, β_1_ = 0.995 ± 0.009, β_2_ = −0.251 ± 0.004, β_age_ = 0.995 ± 0.013, β_agex_ = −0.020 ± 0.000, β_sex_ = 0.501 ± 0.003, and β_city_ = −0.005 ± 0.003. Consequently, we used the constraint of >1000 on cluster number when analyzing the real data as well.

Two specific examples of recovering a simulated dose response are illustrated in **Fig. S2**. One of the examples simulated a dose response which initially increased but then peaked and decreased at high doses (**Fig. S2A**), and the other example simulated an initial slight decrease with dose (a “hormetic” effect) followed by an increase at high doses (**Fig. S2B**). In both situations the simulated dose response shapes were reproduced well by the proposed clustering + CML analysis method.

We performed systematic and detailed simulations with intercepts ranging from −5 to 0, linear dose response terms ranging from −1 to +1 Gy^-1^, and quadratic dose response terms ranging from −1 to +1 Gy^-2^. Numerical results of these simulations for intercepts between −5 and −3, which are more plausible in terms of baseline event occurrence, are described in **Table S1**. The proposed analysis method generally performed well: among >1000 simulated parameter combinations, we found only 4 instances where the proposed modeling approach performed poorly because the 95% CI for the linear and/or quadratic term did not cover the true parameter (**Table S2**).

## SUPPLEMENTARY INFORMATION 2

### Sensitivity Analyses and Refutation Tests using the RERF data Set

#### Refutation Test Methodology

To test the robustness of our causal modeling approach on real RERF data, we first implemented a “synthetic unobserved confounder test”, designed to evaluate how the inclusion of an artificial confounder impacts the estimated ATE. A synthetic variable, referred to as a “synthetic unobserved confounder,” was generated using random samples from a standard normal distribution. This variable was then introduced into the dataset, influencing both the treatment (Dose_Gy) and the outcome (Y) through controlled modifications. The treatment variable was adjusted by multiplying it with an exponential function of the synthetic unobserved confounder, with a strong coefficient of 0.5, while the outcome variable was altered by adding a scaled version of the synthetic unobserved confounder based on a parameter called ‘Confounder_Y_effect’ of various magnitudes and signs (-8 to +8). These modifications simulated a scenario where an unmeasured confounder affects both treatment and outcome, potentially biasing the causal effect estimates.

The modified dataset, including the synthetic unobserved confounder, was then used to fit a new causal forest model. The ATE was calculated for this “confounded” model and compared to the original ATE obtained from the unmodified dataset. By systematically varying the strength of the synthetic unobserved confounder’s effect on the outcome (‘Confounder_Y_effect’) and repeating the experiment 20 times with different random seeds, this test evaluated how sensitive the causal model was to such artificial confounding. The results were visualized using boxplots of confounded ATEs across different levels of ‘Confounder_Y_effect’. This approach provided insights into whether and how much unmeasured confounding could distort causal effect estimates in this analysis.

Next, a noise/weak effect test implemented to assess the robustness of the causal forest model to small, potentially spurious effects. This test introduced a random noise variable into the dataset and examined how it affected the ATE. In this test, a noise variable was generated using random samples from a normal distribution, similar to the synthetic unobserved confounder test. However, unlike the previous test, this noise variable was only used to modify the outcome variable (Y), leaving the treatment variable (Dose_Gy) unchanged. The modification to the outcome was controlled by a parameter called ’weak_effect’, which determined the strength of the noise’s influence on the outcome.

The test created a modified dataset that included this noise variable as an additional feature. A new causal forest model was then fitted to this modified dataset, and the ATE was calculated. This process was repeated multiple times with different random seeds for each level of ’weak_effect’, which took on values of −0.5, 0, and 0.5.

The purpose of this test was to assess whether the model might be overfitting to noise or detecting spurious relationships. If the estimated ATEs from the modified data sets were consistently close to the original ATE, it would suggest that the model was robust to small, random perturbations in the data. Conversely, if the ATEs varied significantly with the introduction of this weak effect, it might indicate that the model was overly sensitive to noise or potential confounding. The results of this test were visualized using boxplots, showing the distribution of ATEs for each level of ’weak_effect’.

Finally, a ‘shuffled treatment + noise test’ was implemented to additionally assess the causal forest model’s ability to detect true causal relationships and resist spurious associations.

This test combined two key elements: randomization of the treatment variable and introduction of noise to the outcome variable.

In this test, a noise variable was generated using random samples from a normal distribution, similar to the previous tests. However, this test went a step further by also randomly shuffling the treatment variable (Dose_Gy). The shuffling process completely broke any existing relationship between the treatment and other variables, including the outcome. The outcome variable was then modified by adding a scaled version of the noise, controlled by the ’weak_effect’ parameter.

The modified dataset included the shuffled treatment, the noise-adjusted outcome, and the noise variable as an additional feature. A new causal forest model was fitted to this modified dataset, and the ATE was calculated. This process was repeated multiple times with different random seeds for each level of ’weak_effect’, which again took on values of −0.5, 0, and 0.5.

Ideally, with a shuffled treatment, the estimated ATE should be close to zero, regardless of the weak effect added to the outcome. Any substantial deviation from zero would suggest that the model might be detecting spurious relationships or overfitting to noise. The results of this test were visualized using boxplots, showing the distribution of ATEs for each level of ’weak_effect’.

#### Refutation Test Results

The results of three such tests are summarized in **Fig. S3**. In the synthetic unobserved confounder test (**Fig. S3A**) the introduced variable was a standard random normal vector (with mean=0 and standard deviation=1). It had an exponential and relatively strong multiplicative effect on the treatment (radiation dose) with a coefficient of 0.5: an exponential effect was used to avoid negative radiation doses. The confounder also had a linear additive effect on the outcome (ln of the mortality event rate) varied from −8 to +8. This range of effects on the outcome varied from very strong negative to very strong positive, including weak and null (0) values in the middle. The null (0 effect) represents a scenario where the synthetic unobserved confounder affects the treatment but has no direct effect on the outcome. This is also an important test case because it shows how the model performs when there is a feature that influences the treatment but not the outcome directly. The results of this test (**Fig. S3A**) suggest that, as expected, the null scenario had no major effect on the average treatment effect (ATE), while increasing the magnitude of the synthetic unobserved confounder effect on the outcome in either direction made the ATE deviate further from the original ATE. The asymmetry in this plot is most likely due to the non-linear (exponential) way the synthetic unobserved confounder affects the treatment, combined with a linear effect on the outcome. Overall, test results suggest that ATE was relatively robust to unmeasured confounding, with strong effects being necessary to perturb it substantially.

In addition to the synthetic unobserved confounder test, a ’noise’ or ’weak effect’ test (**Fig. S3B**) was conducted to assess the sensitivity of the ATE to small, random perturbations in the outcome variable. A standard normal random vector was generated and added to the outcome (natural logarithm of the mortality event rate), scaled by a weak effect parameter that took on values of −0.5, 0, and 0.5. Unlike the synthetic unobserved confounder test, the treatment (radiation dose) was not directly affected in this test. The purpose was to determine if the ATE was susceptible to minor, unmodeled variations in the outcome. As shown in **Fig. S3B**, the ATE remains relatively stable across the different levels of weak effect, with the boxplots for each noise level largely overlapping and centered near the original ATE. This suggests that the estimated causal effect is not overly sensitive to small amounts of random noise in the outcome variable.

Finally, a ’shuffled treatment + noise’ test (**Fig. S3C**) was performed. In this test, the assignment of radiation dose was randomly shuffled across all individuals, breaking any true causal link between dose and mortality. Additionally, as in the ’noise’ test, a small amount of random noise (scaled by values of −0.5, 0, and 0.5) was added to the outcome. The purpose of this test is to confirm that, when the true causal relationship is broken, the model correctly estimates a near-zero ATE. As seen in **Fig. S3C**, the estimated ATEs are indeed centered around zero, with boxplots showing values close to zero for all levels of added noise. This provides confidence that the model does not spuriously detect a treatment effect when none exists.

## Data Availability

https://www.rerf.or.jp/en/library/data-en/

---

**FIG S1.**
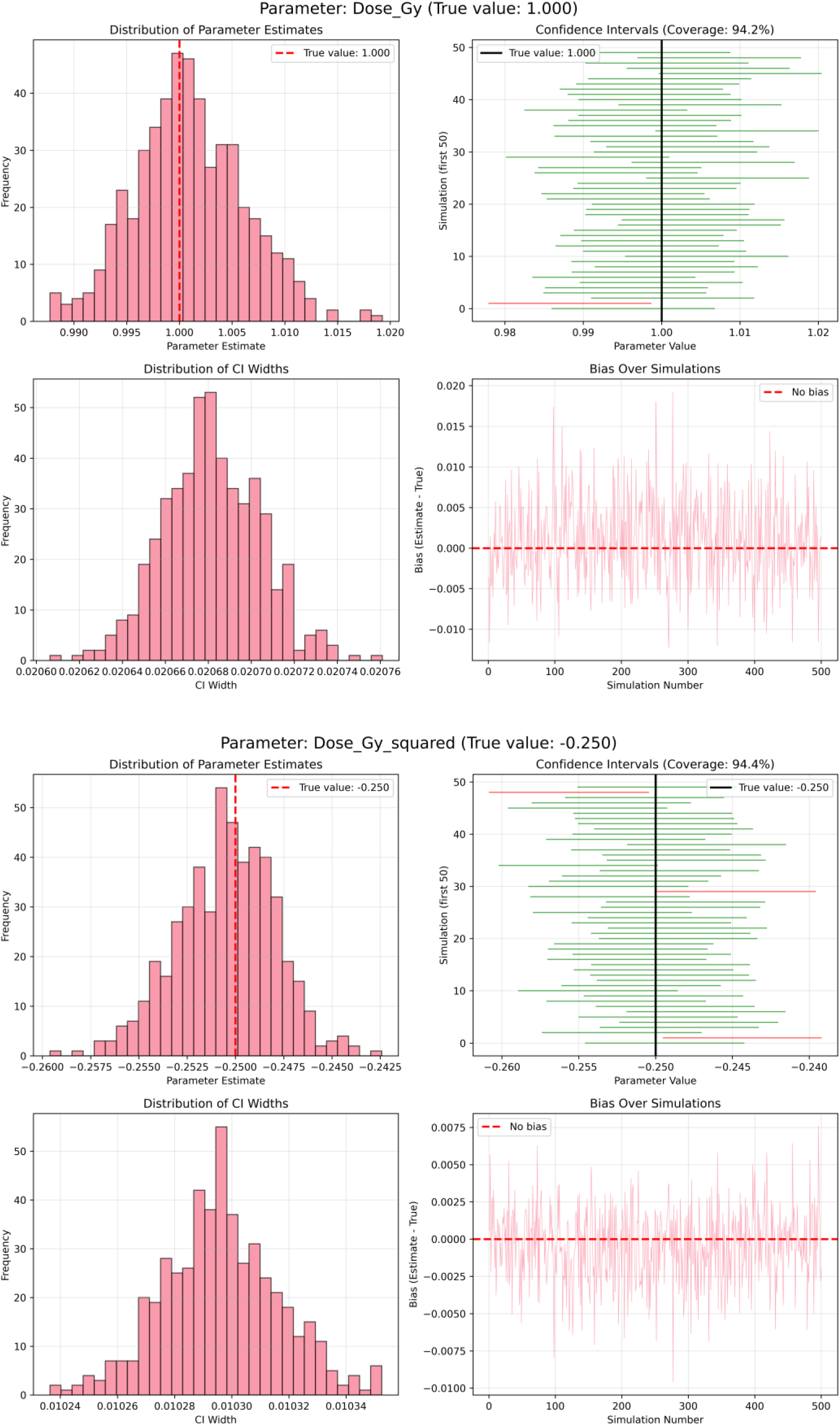
Assessment of the ability of 95% CIs generated by Poisson regression on simulated data after clustering to cover the true parameters of the simulation model. In this example, 500 simulations were used, and the true values for the radiation dose response terms in the data simulation model were α=1.0 Gy^-1^, β=-0.25 Gy^-2^. The top panel shows results for the α (Dose_Gy) parameter and the bottom panel shows results for the β (Dose_Gy_squared) parameter. In both cases, the 95% CI coverages were close to the nominal values: 94.2 and 94.4%, respectively.

**FIG. S2.**
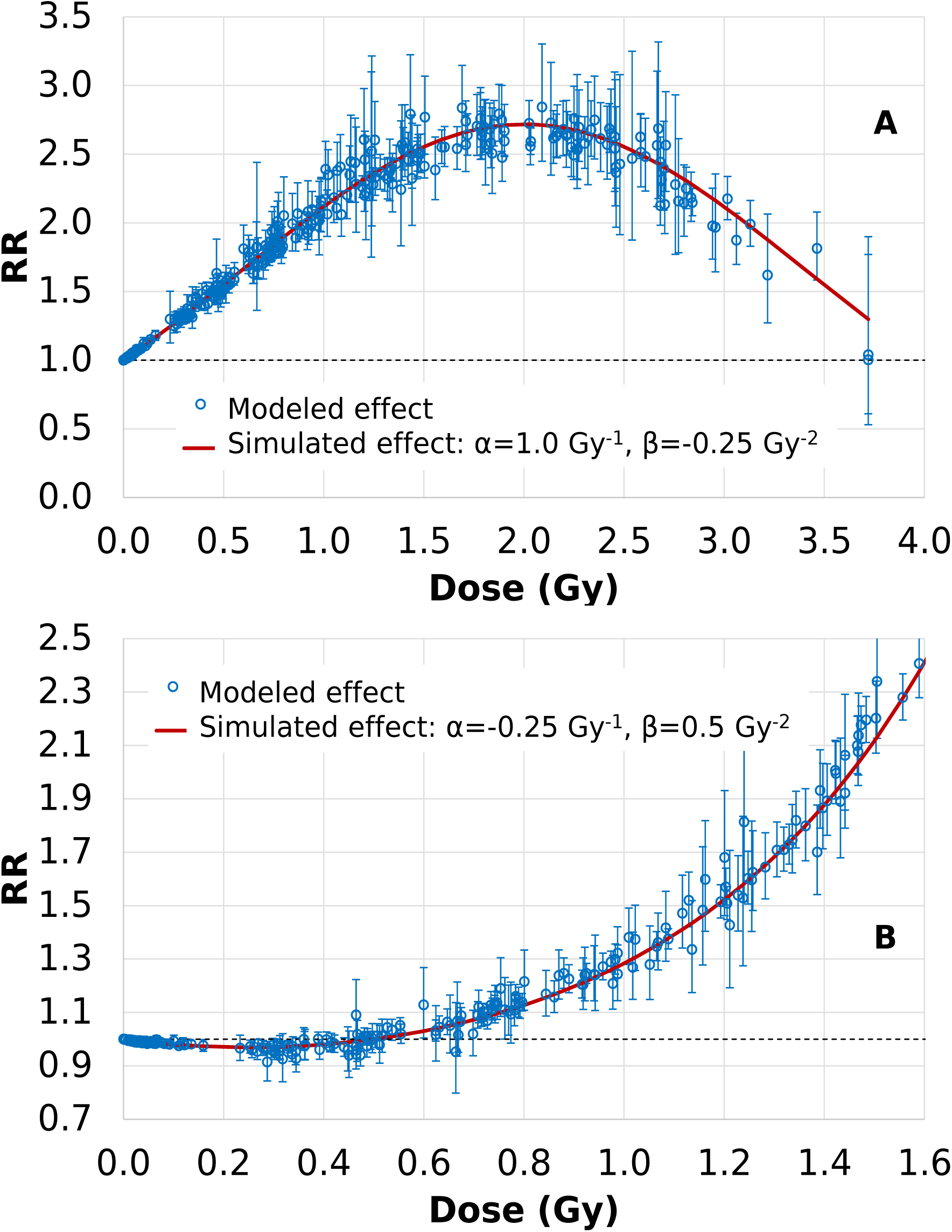
Illustrating the capability of the CML approach used here to reconstruct known simulated dose-response data: Comparison of assumed simulated causal dose-responses (red) *vs*. CML-reconstructed estimates (blue points, with 95% CIs). For illustrative purposes the upper and lower panels correspond to two very different assumed dose-responses (top panel α=1.0 Gy^-1^, β=-0.25 Gy^-2^; bottom panel α=-0.25 Gy^-1^, β=0.5 Gy^-2^). In both panels, coefficients for other features (age, agex, sex and city) were included in the data simulations, but did not play any direct role in the dose response since there were no assumed interaction terms between these features and radiation dose in these simulations.

**FIG. S3.**
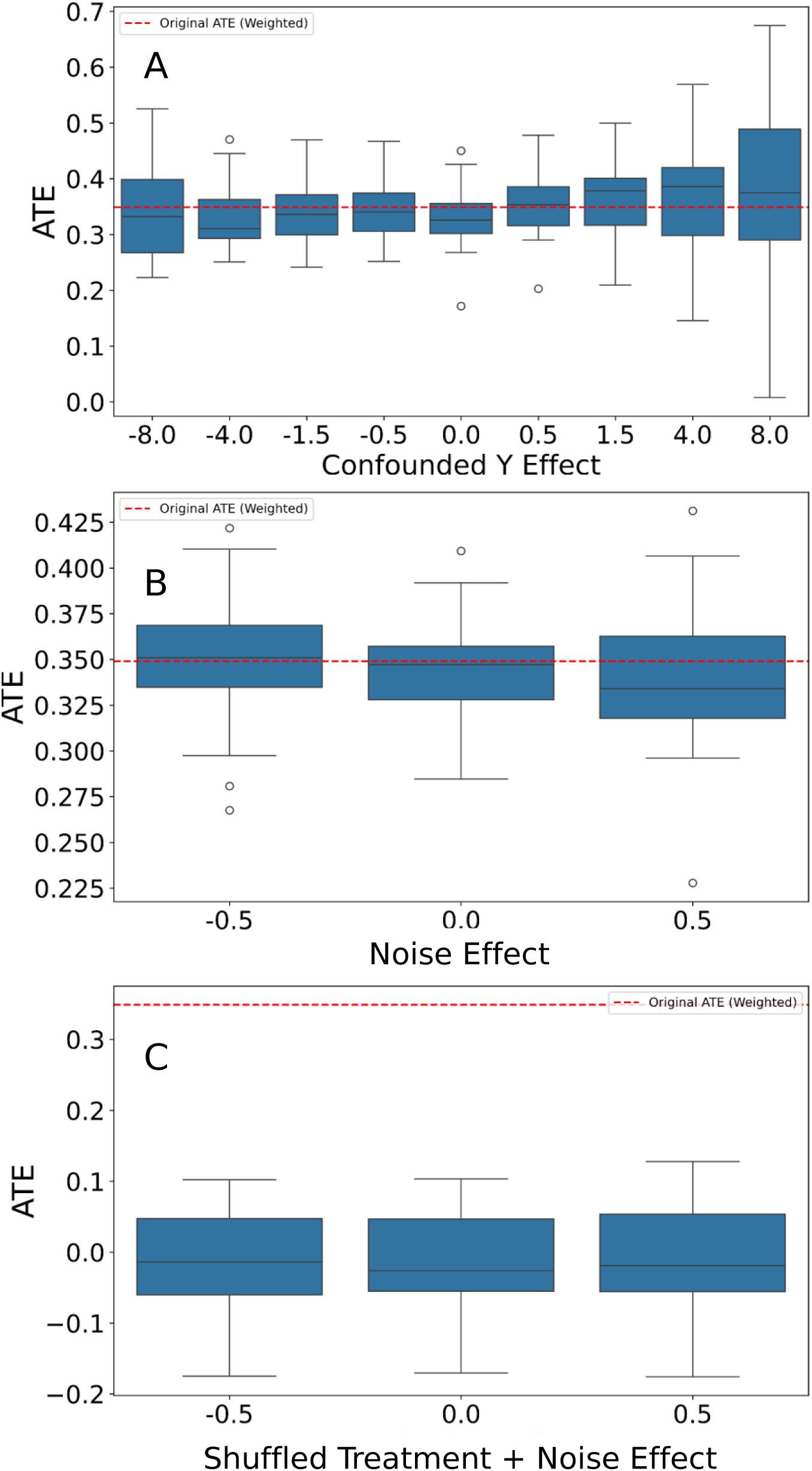
Refutation test results used to validate the robustness of causal radiation effect estimates for all-cause mortality. Panel A = Synthetic unobserved confounder test: A synthetic confounder variable with varying effects on the outcome and treatment is introduced, and the ATE is recalculated multiple times for each confounder effect level. The results are visualized in boxplots to show how the estimated ATE changes with different confounder strengths. Panel B = Noise and weak effect test: Here a synthetic noise variable is added, and it has either zero or small negative or positive effects on the outcome. Panel C = Shuffled treatment + noise effect test: Here the test not only adds random noise to the outcome variable but also randomly shuffles the treatment variable (radiation dose). This test aims to break any potential causal relationship between the treatment and outcome, allowing assessment of the model’s robustness to both noise in the outcome and randomization of the treatment assignment. In all panels the dashed red line indicates the ATE on unperturbed original data.

**TABLE S1.** Performance of the proposed causal forest modeling methodology on various simulated data examples with different dose response parameters. Color coding of columns is included to facilitate comparison of corresponding simulated and modeled parameters.

| Simulated coefficients |  |  | Modeled coefficients from causal forest |  |  |  |  |  |  |  |  |  |
| --- | --- | --- | --- | --- | --- | --- | --- | --- | --- | --- | --- | --- |
| Inter-<br>cept | Line-<br>ar<br>term | Quad-<br>ratic<br>term | Line-<br>ar<br>term | p-value | 95% CI |  | CI<br>covers<br>true<br>para-<br>meter? | Quad-<br>ratic<br>term | p-value | 95% CI |  | CI<br>covers<br>true<br>para-<br>meter? |
| -5.00 | -1.00 | -1.00 | -0.49 | 0.717 | -3.17 | 2.18 | TRUE | -1.33 | 0.539 | -5.56 | 2.91 | TRUE |
| -5.00 | -1.00 | -0.25 | -0.73 | 0.411 | -2.48 | 1.02 | TRUE | -0.39 | 0.789 | -3.21 | 2.44 | TRUE |
| -5.00 | -1.00 | -0.50 | -0.91 | 0.286 | -2.58 | 0.76 | TRUE | -0.35 | 0.780 | -2.80 | 2.10 | TRUE |
| -5.00 | -1.00 | 0.00 | -1.17 | 0.179 | -2.89 | 0.54 | TRUE | 0.07 | 0.952 | -2.11 | 2.25 | TRUE |
| -5.00 | -1.00 | 0.25 | -1.34 | 0.190 | -3.33 | 0.66 | TRUE | 0.48 | 0.283 | -0.40 | 1.36 | TRUE |
| -5.00 | -1.00 | 0.50 | -1.41 | 0.281 | -3.98 | 1.16 | TRUE | 0.72 | 0.146 | -0.25 | 1.68 | TRUE |
| -5.00 | -1.00 | 1.00 | -0.97 | 0.016 | -1.75 | -0.18 | TRUE | 1.01 | 0.000 | 0.64 | 1.38 | TRUE |
| -5.00 | -0.25 | -1.00 | 0.21 | 0.885 | -2.61 | 3.03 | TRUE | -1.35 | 0.534 | -5.62 | 2.91 | TRUE |
| -5.00 | -0.25 | -0.25 | -0.87 | 0.160 | -2.08 | 0.34 | TRUE | 0.23 | 0.687 | -0.87 | 1.33 | TRUE |
| -5.00 | -0.25 | -0.50 | -0.11 | 0.918 | -2.14 | 1.93 | TRUE | -0.76 | 0.604 | -3.61 | 2.10 | TRUE |
| -5.00 | -0.25 | 0.00 | -0.49 | 0.507 | -1.95 | 0.97 | TRUE | 0.18 | 0.551 | -0.42 | 0.79 | TRUE |
| -5.00 | -0.25 | 0.25 | -0.52 | 0.506 | -2.07 | 1.02 | TRUE | 0.39 | 0.190 | -0.20 | 0.98 | TRUE |
| -5.00 | -0.25 | 0.50 | -0.20 | 0.727 | -1.29 | 0.90 | TRUE | 0.49 | 0.061 | -0.02 | 0.99 | TRUE |
| -5.00 | -0.25 | 1.00 | -0.27 | 0.461 | -0.99 | 0.45 | TRUE | 1.03 | 0.000 | 0.68 | 1.38 | TRUE |
| -5.00 | -0.50 | -1.00 | 0.31 | 0.819 | -2.34 | 2.96 | TRUE | -1.87 | 0.371 | -5.97 | 2.23 | TRUE |
| -5.00 | -0.50 | -0.25 | -0.58 | 0.321 | -1.74 | 0.57 | TRUE | -0.09 | 0.906 | -1.51 | 1.33 | TRUE |
| -5.00 | -0.50 | -0.50 | -0.66 | 0.534 | -2.76 | 1.43 | TRUE | -0.42 | 0.789 | -3.53 | 2.68 | TRUE |
| -5.00 | -0.50 | 0.00 | -0.88 | 0.079 | -1.87 | 0.10 | TRUE | 0.32 | 0.329 | -0.32 | 0.96 | TRUE |
| -5.00 | -0.50 | 0.25 | -0.85 | 0.281 | -2.40 | 0.70 | TRUE | 0.43 | 0.130 | -0.13 | 0.99 | TRUE |
| -5.00 | -0.50 | 0.50 | -0.59 | 0.377 | -1.89 | 0.71 | TRUE | 0.56 | 0.046 | 0.01 | 1.12 | TRUE |
| -5.00 | -0.50 | 1.00 | -0.49 | 0.173 | -1.19 | 0.21 | TRUE | 1.01 | 0.000 | 0.68 | 1.33 | TRUE |
| -5.00 | 0.00 | -1.00 | -0.07 | 0.943 | -1.86 | 1.73 | TRUE | -0.84 | 0.569 | -3.72 | 2.04 | TRUE |
| -5.00 | 0.00 | -0.25 | -0.39 | 0.522 | -1.57 | 0.80 | TRUE | -0.14 | 0.843 | -1.52 | 1.24 | TRUE |
| -5.00 | 0.00 | -0.50 | -0.34 | 0.596 | -1.57 | 0.90 | TRUE | -0.18 | 0.853 | -2.09 | 1.73 | TRUE |
| -5.00 | 0.00 | 0.00 | -0.46 | 0.669 | -2.59 | 1.66 | TRUE | 0.27 | 0.456 | -0.44 | 0.98 | TRUE |
| -5.00 | 0.00 | 0.25 | -0.29 | 0.582 | -1.31 | 0.74 | TRUE | 0.40 | 0.096 | -0.07 | 0.86 | TRUE |
| -5.00 | 0.00 | 0.50 | -0.01 | 0.980 | -0.59 | 0.58 | TRUE | 0.52 | 0.000 | 0.26 | 0.79 | TRUE |
| -5.00 | 0.00 | 1.00 | -0.06 | 0.860 | -0.71 | 0.59 | TRUE | 1.06 | 0.000 | 0.72 | 1.40 | TRUE |
| -5.00 | 0.25 | -1.00 | 0.59 | 0.540 | -1.30 | 2.49 | TRUE | -1.31 | 0.360 | -4.11 | 1.49 | TRUE |
| -5.00 | 0.25 | -0.25 | -0.14 | 0.853 | -1.61 | 1.33 | TRUE | -0.02 | 0.957 | -0.65 | 0.62 | TRUE |
| -5.00 | 0.25 | -0.50 | -0.04 | 0.957 | -1.28 | 1.21 | TRUE | -0.41 | 0.595 | -1.91 | 1.09 | TRUE |
| -5.00 | 0.25 | 0.00 | -0.13 | 0.870 | -1.71 | 1.44 | TRUE | 0.23 | 0.494 | -0.43 | 0.89 | TRUE |
| -5.00 | 0.25 | 0.25 | 0.13 | 0.774 | -0.73 | 0.98 | TRUE | 0.32 | 0.108 | -0.07 | 0.72 | TRUE |
| -5.00 | 0.25 | 0.50 | 0.12 | 0.821 | -0.92 | 1.16 | TRUE | 0.56 | 0.035 | 0.04 | 1.09 | TRUE |
| -5.00 | 0.25 | 1.00 | 0.34 | 0.235 | -0.22 | 0.89 | TRUE | 0.99 | 0.000 | 0.67 | 1.30 | TRUE |
| -5.00 | 0.50 | -1.00 | 0.52 | 0.561 | -1.24 | 2.29 | TRUE | -1.05 | 0.426 | -3.64 | 1.54 | TRUE |
| -5.00 | 0.50 | -0.25 | -0.10 | 0.901 | -1.67 | 1.47 | TRUE | 0.09 | 0.748 | -0.45 | 0.63 | TRUE |
| -5.00 | 0.50 | -0.50 | -0.02 | 0.957 | -0.92 | 0.87 | TRUE | -0.14 | 0.801 | -1.26 | 0.98 | TRUE |
| -5.00 | 0.50 | 0.00 | 0.33 | 0.551 | -0.75 | 1.41 | TRUE | 0.09 | 0.700 | -0.38 | 0.57 | TRUE |
| -5.00 | 0.50 | 0.25 | 0.31 | 0.524 | -0.65 | 1.28 | TRUE | 0.32 | 0.167 | -0.14 | 0.78 | TRUE |
| -5.00 | 0.50 | 0.50 | 0.39 | 0.306 | -0.35 | 1.13 | TRUE | 0.57 | 0.004 | 0.18 | 0.95 | TRUE |
| -5.00 | 0.50 | 1.00 | 0.49 | 0.068 | -0.04 | 1.01 | TRUE | 1.05 | 0.000 | 0.77 | 1.33 | TRUE |
| -5.00 | 1.00 | -1.00 | 1.01 | 0.167 | -0.42 | 2.44 | TRUE | -1.09 | 0.275 | -3.04 | 0.86 | TRUE |
| -5.00 | 1.00 | -0.25 | 0.67 | 0.325 | -0.66 | 1.99 | TRUE | -0.08 | 0.795 | -0.67 | 0.51 | TRUE |
| -5.00 | 1.00 | -0.50 | 0.58 | 0.215 | -0.34 | 1.50 | TRUE | -0.30 | 0.331 | -0.89 | 0.30 | TRUE |
| -5.00 | 1.00 | 0.00 | 0.80 | 0.155 | -0.30 | 1.91 | TRUE | 0.09 | 0.743 | -0.43 | 0.61 | TRUE |
| -5.00 | 1.00 | 0.25 | 1.04 | 0.000 | 0.53 | 1.54 | TRUE | 0.26 | 0.044 | 0.01 | 0.51 | TRUE |
| -5.00 | 1.00 | 0.50 | 0.92 | 0.001 | 0.38 | 1.46 | TRUE | 0.55 | 0.000 | 0.29 | 0.81 | TRUE |
| -5.00 | 1.00 | 1.00 | 1.01 | 0.000 | 0.60 | 1.43 | TRUE | 1.05 | 0.000 | 0.86 | 1.23 | TRUE |
| -4.00 | -1.00 | -1.00 | -1.13 | 0.077 | -2.38 | 0.12 | TRUE | -0.52 | 0.530 | -2.13 | 1.10 | TRUE |
| -4.00 | -1.00 | -0.25 | -0.75 | 0.319 | -2.23 | 0.73 | TRUE | -0.42 | 0.681 | -2.39 | 1.56 | TRUE |
| -4.00 | -1.00 | -0.50 | -0.75 | 0.434 | -2.62 | 1.12 | TRUE | -0.67 | 0.579 | -3.04 | 1.70 | TRUE |
| -4.00 | -1.00 | 0.00 | -1.23 | 0.000 | -1.81 | -0.64 | TRUE | 0.24 | 0.505 | -0.47 | 0.95 | TRUE |
| -4.00 | -1.00 | 0.25 | -1.25 | 0.010 | -2.21 | -0.29 | TRUE | 0.35 | 0.241 | -0.23 | 0.92 | TRUE |
| -4.00 | -1.00 | 0.50 | -1.19 | 0.009 | -2.08 | -0.30 | TRUE | 0.57 | 0.000 | 0.25 | 0.89 | TRUE |
| -4.00 | -1.00 | 1.00 | -1.17 | 0.008 | -2.04 | -0.30 | TRUE | 1.09 | 0.000 | 0.67 | 1.50 | TRUE |
| -4.00 | -0.25 | -1.00 | 0.08 | 0.923 | -1.57 | 1.73 | TRUE | -1.26 | 0.225 | -3.30 | 0.78 | TRUE |
| -4.00 | -0.25 | -0.25 | -0.39 | 0.426 | -1.35 | 0.57 | TRUE | -0.18 | 0.765 | -1.38 | 1.02 | TRUE |
| -4.00 | -0.25 | -0.50 | -0.36 | 0.478 | -1.34 | 0.63 | TRUE | -0.33 | 0.591 | -1.53 | 0.87 | TRUE |
| -4.00 | -0.25 | 0.00 | -0.42 | 0.266 | -1.17 | 0.32 | TRUE | 0.08 | 0.710 | -0.35 | 0.51 | TRUE |
| -4.00 | -0.25 | 0.25 | -0.35 | 0.419 | -1.20 | 0.50 | TRUE | 0.29 | 0.094 | -0.05 | 0.64 | TRUE |
| -4.00 | -0.25 | 0.50 | -0.31 | 0.409 | -1.06 | 0.43 | TRUE | 0.53 | 0.002 | 0.20 | 0.86 | TRUE |
| -4.00 | -0.25 | 1.00 | -0.39 | 0.194 | -0.97 | 0.20 | TRUE | 1.08 | 0.000 | 0.79 | 1.37 | TRUE |
| -4.00 | -0.50 | -1.00 | -0.35 | 0.690 | -2.06 | 1.37 | TRUE | -0.99 | 0.362 | -3.13 | 1.14 | TRUE |
| -4.00 | -0.50 | -0.25 | -0.72 | 0.106 | -1.59 | 0.15 | TRUE | -0.08 | 0.893 | -1.23 | 1.08 | TRUE |
| -4.00 | -0.50 | -0.50 | -0.53 | 0.447 | -1.90 | 0.84 | TRUE | -0.42 | 0.619 | -2.10 | 1.25 | TRUE |
| -4.00 | -0.50 | 0.00 | -0.76 | 0.013 | -1.35 | -0.16 | TRUE | 0.16 | 0.529 | -0.34 | 0.66 | TRUE |
| -4.00 | -0.50 | 0.25 | -0.69 | 0.162 | -1.65 | 0.28 | TRUE | 0.31 | 0.082 | -0.04 | 0.67 | TRUE |
| -4.00 | -0.50 | 0.50 | -0.58 | 0.011 | -1.02 | -0.13 | TRUE | 0.53 | 0.000 | 0.33 | 0.72 | TRUE |
| -4.00 | -0.50 | 1.00 | -0.61 | 0.032 | -1.17 | -0.05 | TRUE | 1.06 | 0.000 | 0.79 | 1.32 | TRUE |
| -4.00 | 0.00 | -1.00 | -0.12 | 0.779 | -0.94 | 0.71 | TRUE | -0.79 | 0.083 | -1.69 | 0.10 | TRUE |
| -4.00 | 0.00 | -0.25 | -0.28 | 0.442 | -0.98 | 0.43 | TRUE | -0.15 | 0.734 | -1.00 | 0.70 | TRUE |
| -4.00 | 0.00 | -0.50 | 0.08 | 0.905 | -1.20 | 1.36 | TRUE | -0.61 | 0.419 | -2.07 | 0.86 | TRUE |
| -4.00 | 0.00 | 0.00 | -0.22 | 0.701 | -1.33 | 0.89 | TRUE | 0.07 | 0.691 | -0.29 | 0.43 | TRUE |
| -4.00 | 0.00 | 0.25 | -0.17 | 0.625 | -0.86 | 0.51 | TRUE | 0.31 | 0.056 | -0.01 | 0.63 | TRUE |
| -4.00 | 0.00 | 0.50 | -0.13 | 0.660 | -0.70 | 0.44 | TRUE | 0.56 | 0.000 | 0.28 | 0.84 | TRUE |
| -4.00 | 0.00 | 1.00 | -0.10 | 0.636 | -0.50 | 0.30 | TRUE | 1.07 | 0.000 | 0.91 | 1.23 | TRUE |
| -4.00 | 0.25 | -1.00 | 0.37 | 0.587 | -0.97 | 1.71 | TRUE | -1.07 | 0.154 | -2.54 | 0.40 | TRUE |
| -4.00 | 0.25 | -0.25 | 0.10 | 0.705 | -0.41 | 0.60 | TRUE | -0.22 | 0.458 | -0.79 | 0.36 | TRUE |
| -4.00 | 0.25 | -0.50 | 0.15 | 0.736 | -0.70 | 0.99 | TRUE | -0.45 | 0.366 | -1.43 | 0.53 | TRUE |
| -4.00 | 0.25 | 0.00 | 0.04 | 0.905 | -0.66 | 0.75 | TRUE | 0.09 | 0.546 | -0.20 | 0.38 | TRUE |
| -4.00 | 0.25 | 0.25 | 0.11 | 0.754 | -0.58 | 0.80 | TRUE | 0.31 | 0.054 | -0.01 | 0.63 | TRUE |
| -4.00 | 0.25 | 0.50 | 0.14 | 0.535 | -0.30 | 0.57 | TRUE | 0.56 | 0.000 | 0.36 | 0.76 | TRUE |
| -4.00 | 0.25 | 1.00 | 0.29 | 0.128 | -0.08 | 0.65 | TRUE | 1.02 | 0.000 | 0.86 | 1.19 | TRUE |
| -4.00 | 0.50 | -1.00 | 0.10 | 0.791 | -0.66 | 0.86 | TRUE | -0.57 | 0.163 | -1.37 | 0.23 | TRUE |
| -4.00 | 0.50 | -0.25 | 0.37 | 0.082 | -0.05 | 0.78 | TRUE | -0.19 | 0.226 | -0.49 | 0.12 | TRUE |
| -4.00 | 0.50 | -0.50 | 0.43 | 0.206 | -0.24 | 1.10 | TRUE | -0.51 | 0.231 | -1.33 | 0.32 | TRUE |
| -4.00 | 0.50 | 0.00 | 0.29 | 0.461 | -0.47 | 1.05 | TRUE | 0.08 | 0.641 | -0.24 | 0.40 | TRUE |
| -4.00 | 0.50 | 0.25 | 0.45 | 0.038 | 0.03 | 0.87 | TRUE | 0.27 | 0.009 | 0.07 | 0.48 | TRUE |
| -4.00 | 0.50 | 0.50 | 0.42 | 0.153 | -0.16 | 0.99 | TRUE | 0.55 | 0.000 | 0.26 | 0.84 | TRUE |
| -4.00 | 0.50 | 1.00 | 0.48 | 0.027 | 0.06 | 0.90 | TRUE | 1.04 | 0.000 | 0.84 | 1.24 | TRUE |
| -4.00 | 1.00 | -1.00 | 0.98 | 0.080 | -0.12 | 2.07 | TRUE | -1.02 | 0.086 | -2.18 | 0.14 | TRUE |
| -4.00 | 1.00 | -0.25 | 0.93 | 0.003 | 0.32 | 1.53 | TRUE | -0.23 | 0.055 | -0.47 | 0.01 | TRUE |
| -4.00 | 1.00 | -0.50 | 1.06 | 0.000 | 0.63 | 1.48 | TRUE | -0.56 | 0.009 | -0.98 | -0.14 | TRUE |
| -4.00 | 1.00 | 0.00 | 0.95 | 0.000 | 0.50 | 1.40 | TRUE | 0.03 | 0.788 | -0.22 | 0.29 | TRUE |
| -4.00 | 1.00 | 0.25 | 1.02 | 0.000 | 0.66 | 1.38 | TRUE | 0.25 | 0.005 | 0.07 | 0.42 | TRUE |
| -4.00 | 1.00 | 0.50 | 1.05 | 0.000 | 0.66 | 1.45 | TRUE | 0.50 | 0.000 | 0.31 | 0.69 | TRUE |
| -4.00 | 1.00 | 1.00 | 1.11 | 0.000 | 0.75 | 1.47 | TRUE | 1.01 | 0.000 | 0.84 | 1.17 | TRUE |
| -3.00 | -1.00 | -1.00 | -0.92 | 0.067 | -1.90 | 0.06 | TRUE | -0.93 | 0.128 | -2.13 | 0.27 | TRUE |
| -3.00 | -1.00 | -0.25 | -1.00 | 0.029 | -1.90 | -0.10 | TRUE | -0.18 | 0.722 | -1.16 | 0.81 | TRUE |
| -3.00 | -1.00 | -0.50 | -0.95 | 0.131 | -2.18 | 0.28 | TRUE | -0.49 | 0.507 | -1.96 | 0.97 | TRUE |
| -3.00 | -1.00 | 0.00 | -1.27 | 0.000 | -1.76 | -0.78 | TRUE | 0.20 | 0.455 | -0.32 | 0.72 | TRUE |
| -3.00 | -1.00 | 0.25 | -1.13 | 0.000 | -1.61 | -0.66 | TRUE | 0.33 | 0.001 | 0.14 | 0.52 | TRUE |
| -3.00 | -1.00 | 0.50 | -1.17 | 0.000 | -1.77 | -0.58 | TRUE | 0.58 | 0.000 | 0.35 | 0.81 | TRUE |
| -3.00 | -1.00 | 1.00 | -1.07 | 0.000 | -1.40 | -0.73 | TRUE | 1.05 | 0.000 | 0.87 | 1.22 | TRUE |
| -3.00 | -0.25 | -1.00 | -0.45 | 0.173 | -1.09 | 0.20 | TRUE | -0.68 | 0.049 | -1.35 | 0.00 | TRUE |
| -3.00 | -0.25 | -0.25 | -0.27 | 0.322 | -0.80 | 0.27 | TRUE | -0.22 | 0.423 | -0.76 | 0.32 | TRUE |
| -3.00 | -0.25 | -0.50 | -0.30 | 0.494 | -1.18 | 0.57 | TRUE | -0.41 | 0.410 | -1.37 | 0.56 | TRUE |
| -3.00 | -0.25 | 0.00 | -0.36 | 0.021 | -0.67 | -0.05 | TRUE | 0.03 | 0.812 | -0.23 | 0.29 | TRUE |
| -3.00 | -0.25 | 0.25 | -0.29 | 0.046 | -0.58 | -0.01 | TRUE | 0.26 | 0.000 | 0.13 | 0.38 | TRUE |
| -3.00 | -0.25 | 0.50 | -0.28 | 0.031 | -0.53 | -0.03 | TRUE | 0.51 | 0.000 | 0.39 | 0.64 | TRUE |
| -3.00 | -0.25 | 1.00 | -0.28 | 0.064 | -0.58 | 0.02 | TRUE | 1.04 | 0.000 | 0.92 | 1.16 | TRUE |
| -3.00 | -0.50 | -1.00 | -0.48 | 0.425 | -1.67 | 0.70 | TRUE | -0.94 | 0.204 | -2.40 | 0.51 | TRUE |
| -3.00 | -0.50 | -0.25 | -0.51 | 0.244 | -1.38 | 0.35 | TRUE | -0.27 | 0.573 | -1.21 | 0.67 | TRUE |
| -3.00 | -0.50 | -0.50 | -0.53 | 0.188 | -1.32 | 0.26 | TRUE | -0.51 | 0.247 | -1.37 | 0.35 | TRUE |
| -3.00 | -0.50 | 0.00 | -0.55 | 0.002 | -0.90 | -0.21 | TRUE | 0.03 | 0.859 | -0.30 | 0.36 | TRUE |
| -3.00 | -0.50 | 0.25 | -0.62 | 0.000 | -0.96 | -0.27 | TRUE | 0.29 | 0.000 | 0.15 | 0.43 | TRUE |
| -3.00 | -0.50 | 0.50 | -0.57 | 0.001 | -0.91 | -0.24 | TRUE | 0.53 | 0.000 | 0.38 | 0.68 | TRUE |
| -3.00 | -0.50 | 1.00 | -0.57 | 0.003 | -0.94 | -0.20 | TRUE | 1.04 | 0.000 | 0.86 | 1.23 | TRUE |
| -3.00 | 0.00 | -1.00 | 0.21 | 0.695 | -0.85 | 1.27 | TRUE | -1.13 | 0.067 | -2.34 | 0.08 | TRUE |
| -3.00 | 0.00 | -0.25 | 0.01 | 0.968 | -0.49 | 0.51 | TRUE | -0.25 | 0.341 | -0.78 | 0.27 | TRUE |
| -3.00 | 0.00 | -0.50 | 0.14 | 0.766 | -0.79 | 1.08 | TRUE | -0.62 | 0.219 | -1.60 | 0.37 | TRUE |
| -3.00 | 0.00 | 0.00 | -0.05 | 0.700 | -0.28 | 0.19 | TRUE | 0.00 | 0.958 | -0.14 | 0.14 | TRUE |
| -3.00 | 0.00 | 0.25 | -0.03 | 0.826 | -0.31 | 0.25 | TRUE | 0.27 | 0.000 | 0.14 | 0.40 | TRUE |
| -3.00 | 0.00 | 0.50 | -0.07 | 0.557 | -0.31 | 0.17 | TRUE | 0.53 | 0.000 | 0.42 | 0.64 | TRUE |
| -3.00 | 0.00 | 1.00 | 0.03 | 0.876 | -0.33 | 0.39 | TRUE | 1.01 | 0.000 | 0.85 | 1.17 | TRUE |
| -3.00 | 0.25 | -1.00 | 0.08 | 0.856 | -0.77 | 0.93 | TRUE | -0.77 | 0.105 | -1.70 | 0.16 | TRUE |
| -3.00 | 0.25 | -0.25 | 0.19 | 0.206 | -0.10 | 0.48 | TRUE | -0.23 | 0.055 | -0.46 | 0.01 | TRUE |
| -3.00 | 0.25 | -0.50 | 0.11 | 0.723 | -0.48 | 0.69 | TRUE | -0.39 | 0.154 | -0.94 | 0.15 | TRUE |
| -3.00 | 0.25 | 0.00 | 0.16 | 0.334 | -0.17 | 0.49 | TRUE | 0.03 | 0.667 | -0.12 | 0.18 | TRUE |
| -3.00 | 0.25 | 0.25 | 0.20 | 0.121 | -0.05 | 0.45 | TRUE | 0.27 | 0.000 | 0.15 | 0.39 | TRUE |
| -3.00 | 0.25 | 0.50 | 0.20 | 0.090 | -0.03 | 0.43 | TRUE | 0.53 | 0.000 | 0.43 | 0.63 | TRUE |
| -3.00 | 0.25 | 1.00 | 0.32 | 0.076 | -0.03 | 0.68 | TRUE | 1.01 | 0.000 | 0.87 | 1.16 | TRUE |
| -3.00 | 0.50 | -1.00 | 0.38 | 0.287 | -0.32 | 1.08 | TRUE | -0.86 | 0.018 | -1.58 | -0.15 | TRUE |
| -3.00 | 0.50 | -0.25 | 0.47 | 0.000 | 0.21 | 0.74 | TRUE | -0.27 | 0.018 | -0.49 | -0.05 | TRUE |
| -3.00 | 0.50 | -0.50 | 0.54 | 0.051 | 0.00 | 1.08 | TRUE | -0.53 | 0.038 | -1.03 | -0.03 | TRUE |
| -3.00 | 0.50 | 0.00 | 0.46 | 0.002 | 0.17 | 0.74 | TRUE | 0.01 | 0.918 | -0.13 | 0.15 | TRUE |
| -3.00 | 0.50 | 0.25 | 0.46 | 0.000 | 0.22 | 0.69 | TRUE | 0.26 | 0.000 | 0.16 | 0.37 | TRUE |
| -3.00 | 0.50 | 0.50 | 0.55 | 0.000 | 0.27 | 0.83 | TRUE | 0.49 | 0.000 | 0.37 | 0.61 | TRUE |
| -3.00 | 0.50 | 1.00 | 0.54 | 0.001 | 0.23 | 0.84 | TRUE | 1.02 | 0.000 | 0.90 | 1.14 | TRUE |
| -3.00 | 1.00 | -1.00 | 0.77 | 0.004 | 0.24 | 1.29 | TRUE | -0.78 | 0.001 | -1.26 | -0.30 | TRUE |
| -3.00 | 1.00 | -0.25 | 0.92 | 0.000 | 0.70 | 1.15 | TRUE | -0.24 | 0.000 | -0.34 | -0.13 | TRUE |
| -3.00 | 1.00 | -0.50 | 1.01 | 0.000 | 0.71 | 1.31 | TRUE | -0.52 | 0.000 | -0.77 | -0.27 | TRUE |
| -3.00 | 1.00 | 0.00 | 1.06 | 0.000 | 0.83 | 1.28 | TRUE | -0.03 | 0.594 | -0.13 | 0.07 | TRUE |
| -3.00 | 1.00 | 0.25 | 1.03 | 0.000 | 0.77 | 1.28 | TRUE | 0.25 | 0.000 | 0.14 | 0.35 | TRUE |
| -3.00 | 1.00 | 0.50 | 1.03 | 0.000 | 0.75 | 1.31 | TRUE | 0.51 | 0.000 | 0.39 | 0.62 | TRUE |
| -3.00 | 1.00 | 1.00 | 1.10 | 0.000 | 0.79 | 1.40 | TRUE | 1.00 | 0.000 | 0.87 | 1.13 | TRUE |

**TABLE S2.** Simulated data sets where the proposed causal modeling approach performed poorly because the 95% CI for the linear and/or quadratic term did not cover the true parameter.

| Simulated coefficients |  |  | Modeled coefficients from causal forest |  |  |  |  |  |  |  |  |  |
| --- | --- | --- | --- | --- | --- | --- | --- | --- | --- | --- | --- | --- |
| Inter-<br>cept | Line-<br>ar<br>term | Quad-<br>ratic<br>term | Line-<br>ar<br>term | p-value | 95% CI |  | CI<br>covers<br>true<br>para-<br>meter? | Quad-<br>ratic<br>term | p-value | 95% CI |  | CI<br>covers<br>true<br>para-<br>meter? |
| 0.00 | -1.00 | -1.00 | -1.32 | 0.000 | -1.62 | -1.03 | FALSE | -0.63 | 0.000 | -0.88 | -0.39 | FALSE |
| -0.50 | -0.50 | -1.00 | -0.81 | 0.000 | -1.14 | -0.47 | TRUE | -0.68 | 0.000 | -0.96 | -0.40 | FALSE |
| -1.00 | -0.25 | -1.00 | -0.49 | 0.001 | -0.78 | -0.19 | TRUE | -0.73 | 0.000 | -0.97 | -0.49 | FALSE |

## Notes

### Competing Interest Statement

The authors have declared no competing interest.

### Funding Statement

This study did not receive any specific funding

